# Insulin resistance in thyroid disorders: association between anti-TPO and HOMA-IR

**DOI:** 10.1101/2023.06.06.23291013

**Authors:** Hari Krishnamurthy, Thushani Siriwardhane, Karthik Krishna, Qi Song, Vasanth Jayaraman, Tianhao Wang, Kang Bei, John J. Rajasekaran

## Abstract

The association of thyroid disease and diabetes has been classically described. However, the comorbidity of thyroid disparities and insulin resistance is not frequently assessed, especially the sequence of the occurrence of these markers. We performed a retrospective analysis to evaluate the association between thyroid disease and diabetes markers. We further investigated the sequence of occurrence of thyroid and diabetes markers to identify any predictive capabilities of these markers. We evaluated 32787 subjects who were classified based on their serum thyroid hormones and autoantibody levels. Our general prevalence results showed that HOMA-IR was elevated in overt hypothyroid subjects (43.7%) and overt hyperthyroid subjects (42.2%). HbA1C was elevated in subclinical hypothyroid subjects (19.2%), overt hypothyroid subjects (22.3%) and overt hyperthyroid subjects (21.2%). Glucose was significantly elevated in subclinical hypothyroid subjects (24.2%) and overt hyperthyroid subjects (31.0%). Insulin was only significantly elevated in overt hypothyroid subjects (15.1%). Interestingly, we found that 70.3% of subjects who had their HOMA-IR score escalated from negative (HOMA-IR<2.7) to positive (HOMA-IR>2.7) during their multiple visits had anti-TPO 369 (±242) days prior to the onset of this change. Our comprehensive study provided evidence that the presence of anti-TPO may suggest a predictive role in developing insulin resistance later in life.

**Strengths and limitations of the study:**

- The strength of our study is the large population size including a larger set of markers from both thyroid disease and diabetes.
- The limitation in our study is the distorted male and female ratio.

## INTRODUCTION

Thyroid hormones exert a fine balance in glucose metabolism, acting as both an insulin agonist and an antagonist in different organs. An imbalance, a deficit, or an excess of thyroid hormones could cause disequilibria leading to different disorders, especially in carbohydrate metabolism and insulin resistance.^1^ Thyroid diseases and diabetes are the two most common endocrine disorders encountered in clinical practice. Thyroid disease is mainly classified into hypothyroidism and hyperthyroidism. Elevated levels of thyroid hormones are seen in hyperthyroidism while low levels of thyroid hormones are prevalent in hypothyroidism. Insulin resistance is a condition either caused by defects in insulin production by pancreatic β-cells or defects in effectively employing its function in target cells leading to diabetes. The prevalence of thyroid disease in patients with diabetes is significantly increased than that in the general population. A study conducted by Perros et al. demonstrated a prevalence of 31.4% of thyroid disease in type 1 female diabetes and 6.9% in type 2 male diabetes.^2^ In addition to diabetes, studies have indicated a possible relationship between thyroid status and insulin sensitivity. Though the results of the studies on the effect of thyroid dysfunction on insulin resistance are controversial, many studies have shown that hyperthyroidism may cause impaired glucose tolerance leading to hepatic insulin resistance while hypothyroidism may cause insulin resistance in peripheral tissues.^3^ Additionally, insulin resistance and autoimmune thyroid disease have shown to mutually influence each other. Studies have reported positive connections between thyroid autoantibodies and insulin resistance.^4-6^ Most of these studies have utilized the Homeostatic model assessment (HOMA) method to evaluate the insulin resistance of individuals.

HOMA-IR (HOMA-insulin resistance) is a mathematical model for assessing β-cell function and insulin resistance from fasting glucose and insulin. The HOMA-IR score is determined from glucose and insulin levels in blood in baseline/fasting conditions. In HOMA-IR score, the plasma glucose and insulin concentrations were reflecting the hepatic and peripheral glucose efflux and uptake, while decreases in β-cell function were modeled by changing the β-cell response to plasma glucose concentrations.^7, 8^ The HOMA-IR has proven to be a robust tool for the surrogate assessment of insulin resistance.

Previous studies have suggested a close correlation between insulin resistance and thyroid autoantibodies. However, studies to evaluate the sequence of occurrence of each marker have not been performed. Anti-TPO has been shown to precede years before the onset of many diseases, such as autoimmune thyroid disease, connective tissue disorders, etc.^9-11^ In the present study, we first investigated the prevalence of diabetes markers in thyroid disease individuals and then explored the sequential occurrence of thyroid autoantibodies, anti-TPO, and anti-Tg in relation to HOMA-IR score changes. Understanding the sequential occurrence of these biomarkers will not only help to recognize the disease pathogenesis but also may help for disease prognosis to identify at-risk individuals.

## MATERIALS AND METHODS

### Study Design and Population

A total of 32787 subjects who had suspected thyroid disease or related conditions were tested for thyroid panel and diabetes markers at the Vibrant America Clinical Laboratory by the standard thyroid panel and the diabetes panel between January 2015 and June 2019. Subjects were characterized based on their serum thyroid markers as subclinical hypothyroidism, overt hypothyroidism, subclinical hyperthyroidism, overt hypothyroidism, and controls (thyroid hormones in range). The clinical information on the majority of these patients was provided by physicians as ICD-10-CM (International Classification of Diseases, Tenth Revision, Clinical Modification) codes. The percentage distribution of the top 20 ICD-10-CM codes reported is listed in table S1. This cohort was subdivided into single visit subjects (only visited once between January 2015 and June 2019) and multiple visit subjects (three or more visits between January 2015 and June 2019) for different analysis purposes.

### Thyroid Panel

TSH, FT4, anti-TPO, and anti-Tg were measured using the commercial Roche e601 analyzer (Roche Diagnostics, Indianapolis, IN, USA) according to the manufacturer’s recommendations. All reagents were purchased from Roche Diagnostics (Indianapolis, IN, USA). Human serum specimens were used on Elecsys immunoassay analyzers. Elecsys TSH assay was based on specific TSH monoclonal antibodies specifically directed against human. The antibodies labeled with a ruthenium complex consist of a chimeric construct from human- and mouse-specific components. As a result, interfering effects due to HAMA (human anti-mouse antibodies) were largely eliminated. The Elecsys FT4 test employed a specific anti-T4 antibody labeled with a ruthenium complex to determine the free thyroxine. The Elecsys T3 assay employs polyclonal antibodies specifically directed against T3. Endogenous T3, released by the action of 8-anilino-1- naphthalene sulfonic acid (ANS), competes with the added biotinylated T3-derivative for the binding sites on the antibodies labeled with the ruthenium complex. In the Elecsys FT3 test the determination of free triiodothyronine is made with the aid of a specific anti-T3 antibody labeled with a ruthenium complex. The Elecsys T4 assay employs an antibody specifically directed against T4. Endogenous T4, released by the action of 8-anilino-1-naphthalene sulfonic acid (ANS), competes with the added biotinylated T4-derivative for the binding sites on the antibodies labeled with the ruthenium complex. RT3 was measured using LC-MS on the Xevo TQ-XS mass spectrometer. Elecsys anti-TPO assay employed recombinant antigens and polyclonal anti-TPO antibodies whereas Elecsys anti-Tg assay employed monoclonal human anti-Tg antibodies.

### Reference Ranges for Thyroid Markers

Thyroid hormone reference ranges are subject to the lab where the test is performed. In this study, we followed the reference ranges that majority of the labs used. The reference range of hormones and autoantibody levels in a healthy control used in this study are shown in table 1. Subclinical hypothyroidism, subclinical hyperthyroidism, overt hypothyroidism and overt hyperthyroidism were attributed using these ranges.

**Table 1.** Reference ranges for thyroid markers studied

| Marker | Reference Range |
| --- | --- |
| TSH | 0.3-4.2 mIU/L |
| FT4 | 0.9-1.7 ng/dL |
| Anti- TPO | <9.0 IU/mL |
| Anti-Tg | <4.0 IU/mL |

The categorization of thyroid disease status by evaluating TSH and FT4 levels used in this study is shown in table 2.

**Table 2.** Thyroid disease categorization

| Disease Condition | TSH | FT4 |
| --- | --- | --- |
| Subclinical hypothyroidism | > 4.2 mIU/L | 0.9-1.7 ng/dL |
| Subclinical hyperthyroidism | < 0.3 mIU/L | 0.9-1.7 ng/dL |
| Overt hypothyroidism | > 4.2 mIU/L | < 0.9 ng/dL |
| Overt hyperthyroidism | < 0.3 mIU/L | > 1.7 ng/dL |
| Thyroid Disease negative | 0.3-4.2 mIU/L | 0.9-1.7 ng/dL |

### Diabetes Panel

The Diabetes panel consist of HbA1C, ADIP, Glucose, Glymark, GSP, Insulin. The HbA1c determination was based on the turbidimetric inhibition immunoassay (TINIA) for hemolyzed whole blood. The glucose assay was based on the enzymatic reference method with hexokinase. Hexokinase catalyzed the phosphorylation of glucose to glucose-6-phosphate by ATP. Glucose-6-phosphate dehydrogenase oxidized glucose-6-phosphate in the presence of NADP to gluconate-6-phosphate. No other carbohydrate was oxidized. The rate of NADPH formation during the reaction was directly proportional to the glucose concentration and was measured photometrically. The diazyme insulin assay was used to determine insulin in serum samples. Latex particles coated with antibody specific to human insulin was used to react with insulin present in the sample. Immunoturbidimetry was using Beckman Counter AU series analyzer (Beckman Coulter, Inc., Georgia, USA) was used to measure the turbidity at 600nm primary and 800 nm secondary. The Glycated serum protein assay used proteinase K to digest GSP into low molecular weight glycated protein fragments and used diazyme’s specific fructosaminase. The colorimetric product was measured at 546-600 nm. ADIP assay was based on adiponectin and anti-adiponectin coated latex complexation. The turbidity of the antibody-antigen complex was measured at 570 nm via Roche Cobas 6000 c501 (Roche Diagnostics, Indiana, USA).

### HOMA-IR Calculation

HOMA-IR was calculated as:

HOMA-IR = [Fasting Plasma Glucose (mg/dl) × Fasting Insulin(μU/ml)]/405

### Patient and Public Involvement

This study does not include any patient or public involvement and is based on retrospective analysis of de-identified laboratory data. IRB exemption (work order #1-1098539-1) was determined by the Western Institutional Review Board (WIRB) for Vibrant America Biorepository to use de-linked and de-identified remnant human specimen and medical data for research purposes.

### Statistical Analysis

The data were analyzed using Java for Windows version 1.8.161 and R for Windows version 3.5.0. Data were expressed as mean□±□standard deviation (SD). The association between PSA and autoimmune markers and controls were tested using nonparametric chi-square test with 95% confidential interval. P value < 0.05 was considered statistically significant.

## RESULTS

### Patient Clinical Characteristics

A total of 32787 subjects who ordered the thyroid panel and the diabetes panel were included in this retrospective study. Table 3 shows the demographics of thyroid positive subjects (different thyroid diseases based on their serum thyroid hormone levels) and negative controls in this study.

**Table 3.** Demographics of subjects with different thyroid disease profiles.

|  | Subclinical Hypothyroidism | Overt Hypothyroidism | Subclinical Hyperthyroidism | Overt Hyperthyroidism | Thyroid- (Control) |
| --- | --- | --- | --- | --- | --- |
| N | 3455/52662<br>(6.6%) | 637/52662<br>(1.2%) | 1626/52662<br>(3.1%) | 494/52662<br>(0.9%) | 46450/52662<br>(88.2) |
| Gender | 1258M/2197F | 134M/503F | 168M/1458F | 72M/422F | 16812M/29638F |
| Age | 50.0M/48.2F | 53.4M/51.3F | 55.8M/52.5F | 52.9M/53.0F | 47.1M/45.6F |

### Prevalence of Diabetes markers in thyroid disease

To evaluate the general prevalence of diabetes markers in thyroid disease subjects, we first assessed single visit subjects who had tested for both thyroid disease and diabetes markers (HOMA-IR, HbA1C, ADIP, Glucose, Glymark, GSP, Insulin). Figure 1 shows the diabetes markers significantly elevated in different thyroid disease subjects grouped as subclinical hypothyroidism, overt hypothyroidism, subclinical hyperthyroidism, overt hyperthyroidism based on their serum TSH and FT4 levels. HOMA-IR was elevated in overt hypothyroid subjects (171/391, 43.7%) and overt hyperthyroid subjects (129/306, 42.2%) compared to its controls, the thyroid negative group (10763/28896, 37.2%). HbA1C was elevated in subclinical hypothyroid subjects (639/3325, 19.2%), overt hypothyroid subjects (137/614, 22.3%) and overt hyperthyroid subjects (101/476, 21.2%) compared to its controls (7237/44829, 16.1%). Glucose was significantly elevated in subclinical hypothyroid subjects (579/2394, 24.2%) and overt hyperthyroid subjects (106/342, 31.0%) compared to its control (6984/31803, 22%). Insulin was only significantly elevated in overt hypothyroid subjects (72/478, 15.1%) compared to its control (4022/35197, 11.4%). Subclinical hyperthyroidism did not a show significant relationship with any of the diabetes markers.

**Figure 1.**
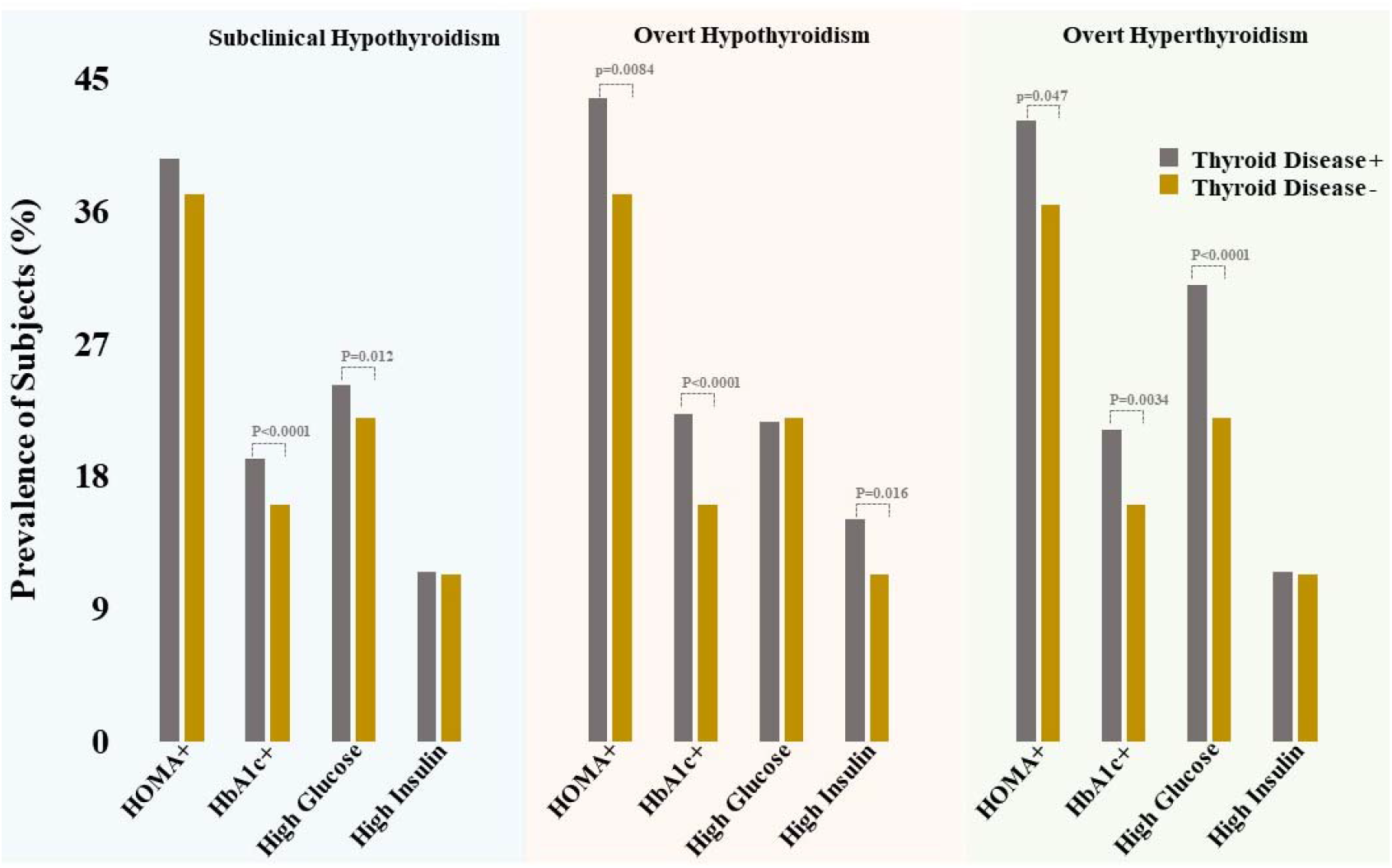
Prevalence of diabetes markers in subjects with different thyroid disease profiles compared to their controls (thyroid disease negative). Diabetes markers significantly elevated/reduced are only shown.

Next, we tested the prevalence of these diabetes markers in antibodies (Anti TPO, Anti Tg) and individual thyroid hormones (TSH, FT4, FT3, RT3, T3, T4). Table 4 provides the prevalence of diabetes markers in subjects with elevated thyroid autoantibodies. HOMA-IR score was elevated in subjects seropositive to anti-TPO and anti-Tg. High insulin levels were seen in subjects with anti-TPO positivity. The prevalence of diabetes markers in subjects with elevated/reduced levels of individual thyroid hormones are listed in table S2.

**Table 4.** Prevalence of diabetes markers in subjects with elevated thyroid autoantibodies.

|  | Anti TPO+ | Anti TPO- | P Value | Anti Tg+ | Anti Tg- | P Value |
| --- | --- | --- | --- | --- | --- | --- |
| HOMA-IR Positive | **45.5%**<br>**(1869/4104)** | **37.5%**<br>**(11015/29387)** | **<0.0001** | **41.1%**<br>**(1747/4255)** | **37%**<br>**(8285/22364)** | **<0.0001** |
| High HbA1C | 16.3%<br>(990/6076) | 15.4%<br>(6502/42088) | 0.0930 | 14%<br>(872/6210) | 14.7%<br>(4701/32057) | 0.2100 |
| Low ADIP | 2.2% (94/4184) | 3.1% (918/29592) | 0.0028 | 1.7% (74/4391) | 3% (671/22699) | <0.0001 |
| High Glucose | 22.1%<br>(1001/4525) | 22%<br>(7097/32218) | 0.9022 | 19.8%<br>(908/4595) | 21.5%<br>(5186/24139) | 0.0093 |
| Low Glucose | 1.7% (75/4525) | 1.8% (567/32218) | 0.6659 | 1.8% (84/4595) | 1.9%<br>(463/24139) | 0.7262 |
| Low Glymark | 10.3%<br>(314/3058) | 10.1%<br>(2250/22345) | 0.7564 | 8.6%<br>(259/2998) | 10.1%<br>(1509/14881) | 0.0132 |
| GSP | 10.7%<br>(438/4104) | 10.7%<br>(3159/29601) | 1.0000 | 11.3%<br>(476/4215) | 11.4%<br>(2540/22266) | 0.8507 |
| High Insulin | **14.1%**<br>**(266/1888)** | **11.5%**<br>**(4490/39169)** | **0.0006** | 11.2%<br>(588/5238) | 11.1%<br>(2990/26991) | 0.7735 |
| Low Insulin | 2.9%<br>(144/5005) | 2.8% (986/34815) | 0.8936 | 2.8%<br>(149/5238) | 2.8%<br>(764/26991) | 0.9916 |

### Early detection of anti-TPO antibodies predicted HOMA-IR score change

To evaluate the direction of disease correlation between diabetes and thyroid disease we extracted subjects who had multiple visits during the time of the study (January 2015 to June 2019). First, a sub cohort who had HOMA-IR<2.7 (negative) in their initial test but increased over time (HOMA-IR>2.7-positive) during their subsequent visits were selected to analyze their clinical profiles to identify any predictive markers for insulin resistance. A total of 155 subjects was included in this group. As shown in figure 2, 109/155 (70.3%) had anti-TPO 369 (± 242) days (ranging from 55 days to 1229 days) prior to the onset of HOMA-IR change from negative to positive. It was significantly different from control group which had 46/155 (29.7%) subjects with either no anti-TPO presence or anti-TPO on or after the negative HOMA-IR change to positivity. Anti-Tg (56/155, 36.1%) or any thyroid disease profile (subclinical hypothyroidism-8/155, 5.2%, overt hypothyroidism-2/155, 1.3%, subclinical hyperthyroidism-12/155, 7.7%, overt hypothyroidism-2/155, 1.3%) did not show this predictive behavior. Next, we analyzed the early occurrence of HOMA-IR positivity in subjects who have converted from seronegative to seropositive for anti-TPO, anti-Tg and thyroid profiles to identify whether high HOMA-IR scores could predict thyroid disease. Of 206 subjects, 36 (17.5%) had HOMA-IR positivity before the onset of anti-TPO seropositivity. Moreover, 13/70 (18.6%) subjects only had HOMA-IR positivity before the onset of thyroid disease and 16/64 (25.0%) had HOMA-IR change prior to the onset of anti-Tg negativity. These data did not show any significant relationship with HOMA-IR positivity occurring ahead of either anti-TPO, anti-Tg or thyroid disease.

**Figure 2.**
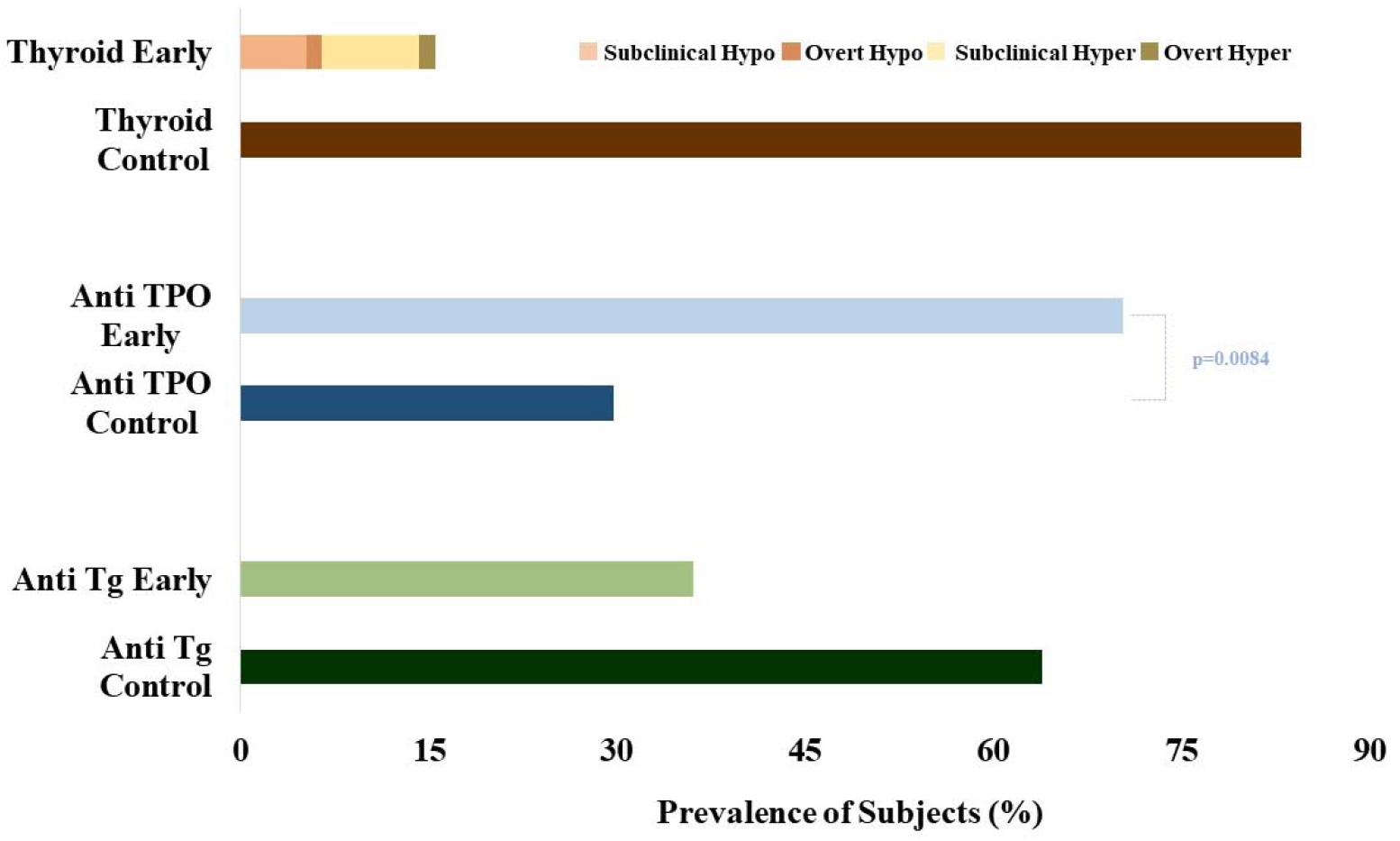
The prevalence of different thyroid diseases, anti-TPO, and anti-Tg prior to the onset of HOMA-IR elevation compared to their respective control groups. Thyroid control group consists of subjects who had thyroid disease-parallel, thyroid disease-following, and no thyroid disease to the onset of HOMA elevation. Anti-TPO control group consists of subjects who had anti-TPO-parallel, anti-TPO -following, and no anti-TPO to the onset of HOMA elevation. Anti-Tg control group consists of subjects who had anti-Tg-parallel, anti-Tg-following, and no anti-Tg to the onset of HOMA elevation.

## DISCUSSION

Our study is the first in the literature evaluating the sequence of markers occurring in thyroid disease and insulin resistance comorbidity. Patients with thyroid disparities are known to have glucose and insulin metabolism disorders, however, none of the studies have elucidated the disease marker sequence. The direction of disease correlation is important for effective follow-up testing and treatment strategies for at-risk subjects.

To understand the general prevalence, we first evaluated the association between thyroid disease and diabetes markers in single visit subjects. We evaluated thyroid disease profiles categorized based on their serum TSH and FT4 levels. We found that both overt hypothyroidism and overt hyperthyroidism had elevated levels of HOMA-IR. HbA1C was elevated in subclinical hypothyroidism, overt hypothyroidism and overt hyperthyroidism. Glucose was elevated in both subclinical hypothyroidism and overt hyperthyroidism and insulin is only elevated in overt hypothyroidism. Clinical hyperthyroidism is associated with abnormal glucose tolerance and insulin resistance.^12^ Specifically, overt hyperthyroidism has increased demand for insulin which is often due to accelerated metabolism, tissue resistance to insulin, and elevated insulin degradation.^3^ Many theories have established to explain the peripheral insulin resistance in hyperthyroidism. Some studies show that increased secretion of adiponectin such as interleukin-6, and TNF- α in hyperthyroid patients may be related to the development of insulin resistance.^14^ Another mechanism by which thyroid hormones increase hepatic glucose production is through the increased expression of GLUT2 transporter on hepatic plasma membrane which was found to be twice as high in hyperthyroid patients compared to hypothyroid patients.^1^ There are comparatively lesser human studies conducted to evaluate the association between hypothyroidism and insulin resistance. In hypothyroidism, it is speculated that decreased intestinal glucose absorption rate and decreased adrenergic activity may lead to a reduction in glycogenolysis in the liver and muscles and gluconeogenesis that could further decrease the baseline insulin secretion.^12, 15^ Our results indicated a significant prevalence of increased HOMA-IR scores in hypothyroid subjects indicating a possible insulin resistance. Similar results were also seen by other groups where they found hypothyroidism induced a decrease in the insulin-mediated glucose disposal that reverted upon treatment.^16-18^

Autoimmune thyroid disease and insulin resistance are two chronic inflammatory diseases. Hence, we evaluated the relationship between subjects that were seropositive for thyroid autoantibodies and insulin resistance. Our results showed that subjects with thyroid antibodies, anti-TPO and anti-Tg had significantly elevated HOMA-IR scores compared to their respective control groups. Moreover, seropositive subjects for anti-TPO had elevated insulin levels compared to seronegative subjects. Studies have shown a connection between anti-TPO titer, the secretory function of monocytes, lymphocytes and pro-inflammatory cytokines such as TNF-α and IL-6.^19^ These monocytes, lymphocytes and their secretary cytokines TNF-α and IL-6 are also responsible for developing insulin resistance.^20^ Our results were consistent with Tina Mazaheri et al. where they found significantly high insulin levels in subjects with anti TPO antibodies more than 1000 IU/ml.^21^ However, their HOMA-IR score did not show any significant difference.

It has long been a topic of controversy about the direction of the effect of insulin resistance and autoimmune thyroid disease on each other. To our knowledge, none of the studies have evaluated the direction of these two comorbid diseases, diabetes and thyroid disease. To assess the sequence of occurrence of these disease markers, we evaluated the predictive characteristics of anti-TPO, anti Tg, and thyroid disease in insulin resistance and vice versa. Interestingly, we found that 70.3% subjects had anti-TPO prior to the HOMA-IR change from negative to positive, indicating a possible predictive role of anti-TPO in insulin resistance. None of the thyroid disease profiles (15.5%) nor anti-Tg (36.1%) preceded the HOMA-IR score change, indicating that thyroid disease profiles or anti-Tg does not contain any significant predictability on insulin resistance. However, the opposite direction, the appearance of insulin resistance, in terms of high HOMA-IR scores, appearing before anti-TPO positivity, anti-Tg positivity or any thyroid disease positivity did not provide any significant results, eliminating the hypothesis of the occurrence of insulin resistance before anti-TPO, anti-Tg or thyroid disease seropositivity.

In this study, we mainly examined the effect of direction of comorbid diseases, thyroid disease and insulin resistance. The primary strength of our study is the large population size including a larger set of markers from both thyroid disease and diabetes. However, a limitation in our study is the distorted male and female ratio. A study with similar male and female ratio with all-inclusive clinical characteristics such as thyroid treatments, female menopausal status, etc. would be required to extrapolate the results to eventually achieve a generalized outcome. In conclusion, we showcased that thyroid autoantibody, anti-TPO precedes the changes in insulin resistance in terms of HOMA-IR score changes from negative to positive, thus anti-TPO could eventually be used as a predictive marker for disease stratification.

## Data Availability

The raw data supporting the conclusions of this manuscript will be made available by the corresponding author on reasonable request.

## Consent for Publication

N/A

## Author contributions

Conception and study design: HK, JR, VJ. Performing experiments: KK, TW. Analysis and interpretation: TS, KB, QS, HK. Manuscript preparation: TS, HK. All authors read and approved the final manuscript.

## Acknowledgement

We acknowledge Vibrant America LLC for supporting this research.

## Ethics approval and consent to participate

The study was conducted under the ethical principles that have their origins in the Declaration of Helsinki. This is a retrospective study conducted from remnant samples of individuals that visited Vibrant Clinical Lab for routine tests. The IRB (WCG IRB (Western Institutional Review Board) (#1-1098539-1)) exemption enabled us to use the de-identified laboratory data for the retrospective analysis. The data and materials in this manuscript have not been published elsewhere and are not under consideration by another journal.

