## Supplementary table for "Insulin resistance in thyroid disorders: association between anti-TPO and HOMA-IR": SI Insulin resistance in thyroid disorders association between anti-TPO and HOMA-IR.docx

**Supplementary Material**

Table S1. Top 20 physician-reported ICD-10-CM codes distribution for subjects tested for thyroid disease.

| **ICD Code** | **Description** | **Hypothyroidism (%)** | **Hyperthyroidism (%)** |
| --- | --- | --- | --- |
| R5383 | Other fatigue | 51.0 | 55.7 |
| E559 | Vitamin D deficiency, unspecified | 33.3 | 41.2 |
| E039 | Hypothyroidism, unspecified | 24.9 | 41.5 |
| M2550 | Pain in unspecified joint | 18.5 | 13.0 |
| E782 | Mixed hyperlipidemia | 13.1 | 16.7 |
| I10 | Essential (primary) hypertension | 11.3 | 8.1 |
| E785 | Hyperlipidemia, unspecified | 10.7 | 10.0 |
| K5900 | Constipation, unspecified | 9.1 | 7.9 |
| E349 | Endocrine disorder, unspecified | 8.2 | 13.5 |
| K909 | Intestinal malabsorption, unspecified | 7.6 | 5.6 |
| D539 | Nutritional anemia, unspecified | 7.5 | 8.4 |
| R7989 | Other specified abnormal findings of blood chemistry | 6.8 | 7.4 |
| Z79899 | Other long term (current) drug therapy | 6.6 | 8.9 |
| R638 | Other symptoms and signs concerning food and fluid intake | 6.0 | 5.5 |
| R5382 | Chronic fatigue, unspecified | 5.5 | 5.1 |
| N951 | Menopausal and female climacteric states | 5.3 | 17.5 |
| R531 | Weakness | 5.1 | 2.9 |
| R5381 | Other malaise | 4.8 | 3.8 |
| E663 | Overweight | 4.7 | 2.2 |
| Z0000 | Encounter for general adult medical examination without abnormal findings | 4.7 | 5.0 |

Table S2. Prevalence of diabetes markers in subjects with elevated/reduced levels of other thyroid hormones. Bold percentages are significantly different (p<0.05) compared to their respective control groups.

|  | Elevated FT3 | Reduced FT3 | FT3 in range | Elevated T4 | Reduced T4 | T4 in range | Elevated T3 | Reduced T3 | T3 in range | Elevated RT3 | Reduced RT3 | RT3 in range |
| --- | --- | --- | --- | --- | --- | --- | --- | --- | --- | --- | --- | --- |
| HOMA-IR | **41.1% (500/1217)** | 33.2% (87/262) | **37.7% (12623/33523)** | **47.6% (709/1490)** | 33.3% (240/720) | **37.7% (11366/30168)** | **49.2% (229/465)** | 23.2% (257/1108) | **38.4% (11649/30365)** | **41.3% (1054/2549)** | 39.1% (361/924) | **36.8% (9544/25955)** |
| HbA1C | 12.2% (244/2003) | 21.2% (86/406) | 16.3% (8553/52368) | **21.3% (424/1989)** | 13.1% (132/1008) | **16% (6658/41521)** | 15.9% (92/577) | 14.2% (211/1490) | 16.1% (6625/41081) | **25.4% (869/3428)** | 12.5% (166/1330) | **14.5% (5413/37302)** |
| Low ADIP | **3% (35/1154)** | 0.8% (2/259) | **2% (671/33399)** | 2.3% (35/1528) | 4.5% (32/709) | 3% (888/29787) | **5.3% (24/453)** | 1.9% (20/1063) | **3% (900/30149)** | 2.5% (67/2673) | 3.5% (33/940) | 2.8% (722/26162) |
| High Glucose | 21.8% (297/1362) | **28% (82/293)** | 22.3% (8240/36915) | 23.5% (393/1670) | 25.2% (196/778) | 22.2% (7392/33238) | 24.2% (123/508) | 20.1% (245/1217) | 22.3% (7427/33373) | **26.4% (745/2826)** | 20.5% (209/1018) | **21.2% (5968/28198)** |
| Low Glucose | 1.8% (25/1362) | 3.1% (9/293) | 1.7% (632/36915) | **2.4% (40/1670)** | 1.9% (15/778) | **1.7% (549/33238)** | 2.4% (12/508) | 2.3% (28/1217) | 1.7% (555/33373) | **2.5% (70/2826)** | 2.2% (22/1018) | **1.8% (505/28198)** |
| Low Glymark | 7.7% (70/906) | **28.5% (59/207)** | 10% (2505/25025) | 10.2% (126/1234) | 12.2% (70/572) | 10% (2356/23451) | 6.7% (26/390) | **20.2% (165/815)** | 9.8% (2334/23752) | 11.9% (289/2423) | 11.8% (82/696) | 9.7% (1729/17802) |
| GSP | 5.9% (70/1186) | **27.1% (74/273)** | 10.7% (3549/33260) | 11.1% (174/1565) | 11.9% (87/732) | 10.7% (3292/30863) | 6.3% (30/477) | 20.5% (233/1139) | 10.5% (3257/31143) | **13.2% (357/2698)** | 10.1% (95/941) | **10.9% (2827/25893)** |
| High Insulin | 11.6% (175/1508) | 9.8% (31/316) | 11.5% (4747/41458) | 12.1% (201/1657) | 10.8% (90/830) | **10.9% (3736/34158)** | **16.9% (85/504)** | 7.3% (91/1248) | **11.3% (3830/33971)** | 12.2% (343/2805) | 13.2% (146/1110) | 11.3% (3561/31547) |
| Low Insulin | 2.9% (43/1508) | **14.2% (45/316)** | 2.8% (1146/41458) | 1.8% (30/1657) | 3.6% (30/830) | 2.9% (987/34158) | 2.4% (12/504) | **9.9% (124/1248)** | 2.6% (892/33971) | **3.6% (102/2805)** | 3.2% (36/1110) | **2.8% (876/31547)** |
